# Early and accurate prediction of COVID-19 hospitalization risk and symptomatic course of SARS-CoV-2 infection

**DOI:** 10.1101/2024.07.05.24309641

**Authors:** Corinna Holetschek, Mehmet Goekkaya, Karim Dorgham, Paul Eder, Daria Luschkova, Christophe Parizot, Mehmet Tekinsoy, Denise Rauer, Assia Samri, Early-Opt-COVID19 study group, Matthias Reiger, Gertrud Hammel, Philipp Steininger, Stefanie Gilles, Ulrike Protzer, Christoph Römmele, Guy Gorochov, Claudia Traidl-Hoffmann, Avidan U. Neumann

## Abstract

**Background:** Whilst SARS-CoV-2 infection has become endemic, COVID-19 related hospitalization and mortality are still considerably high. Both anti-viral and immune modulating therapies against COVID-19 are available, but they must be initiated early after infection and given only to patients of need. Currently, patients’ demographics and clinical pre-conditions factors are used to determine treatment eligibility. However, the latter do not provide accurate prediction and there are no useful biomarkers for early accurate prediction of COVID-19 related hospitalization risk and disease progression.

**Methods:** Non-vaccinated patients (N=185) were recruited early after the first positive SARS-CoV-2 test. Biochemistry, hematology and 8 serum cytokine levels were longitudinally measured within the first month.

**Findings:** Early levels of LDH, IL-6 or CRP, each alone or their combinations, were identified as accurate predictors for the risk of hospitalization (sensitivity=93.6-100%, specificity=93.4-96.7%, p<0.0001). Moreover, the combination of 4 cytokines (IFN-α, IFN-γ, IL-6, IL-17A) was the only accurate predictor for symptoms risk (sensitivity=97.5%, specificity=92.3%, p<0.0001). In comparison, age and BMI showed significantly lower predictive values than above biomarkers. Prediction with above biomarkers was independent of sampling time (0-11 days post symptoms onset), age, gender, BMI, clinical pre-conditions or SARS-CoV-2 variant. Furthermore, the early higher levels of LDH, CRP and inflammatory cytokines in hospitalized, as compared to non-hospitalized, patients, stayed consistently higher for at least 4 weeks.

**Interpretation:** The risk for COVID-19 hospitalization or symptoms can be accurately predicted as early as the time of the first positive SARS-CoV-2 test, with biomarkers that are feasibly measurable at point-of-testing. These findings could allow for better early personalized treatment and optimization of clinical management of COVID-19 patients.

## Introduction

The SARS-CoV-2 virus started spreading in December 2019 causing a total of 6 million COVID-19 related deaths by December 31, 2021^1^. In the last 2 years, SARS-CoV-2 became endemic, but COVID-19 mortality is still considerably high, with about 200,000 deaths in 2023, even with the less severe Omicron variant^2^. Several Anti-viral drugs are available^3,4,5^ and more are under investigation^6,7^, also for patients with mild-moderate disease. Howevere, these need to be initiated early within 5-7 days post symptoms onset (DPSO) and targeted to patients that are of risk for hospitalization or develop considerable symptomatic disease. Also, immune modulating drugs that inhibit inflammatory cytokines elevated as part of the COVID-19 cytokine storm (e.g., IL-6 inhibitors) are available^8,9^. However, currently, anti-viral therapy is underused^10^ and cytokine inhibitors are only given to hospitalized severe patients, mainly because of lack of biomarkers that allow early stratification of the appropriate target patients^11^. Therefore, it is important to understand which SARS-CoV-2 infected individuals are at risk for hospitalization and/or to suffer from symptomatic mild to moderate disease, in order to allow early personalized treatment and optimize public health management.

COVID-19 shows a versatile range of severity, from asymptomatic, to symptomatic mild-moderate disease and up to critical severity leading to hospitalization, requiring invasive oxygen support, and potentially leading to multi-organ failure and death^12^. COVID-19 has a mean incubation period 5.2 days (95% CI: 4.9–5.5) until symptoms onset^13^. Common early symptoms are characterized by fever, cough, fatigue, diarrhea, and dyspnea^14^. However, meta-analysis showed that 40-50% of confirmed SARS-CoV-2 positive cases are asymptomatic^15^. The Kings College large app study identified the early symptoms that are most associated with subsequent hospitalization and thus allowed stratification of patients into low versus high symptomatic groups^16^.

Patients’ characteristics, such as age and BMI, and underlying comorbidities (e.g., diabetes, asthma, cancer, cardiovascular disorder, chronic kidney disease) are known factors associated with risk for severe COVID-19 disease^17^ and are currently used as recommendations for early treatment with anti-viral therapy^18^. However, these factors do not allow accurate prediction of hospitalization and symptomatic risk. In addition, a number of biochemistry and hematological blood biomarkers were reported to be risk factors for severe COVID-19 in hospitalized patients. C-reactive protein (CRP) levels were found to be significantly higher in severe hospitalized COVID-19 compared to moderate cases^19^, and elevated blood lactate dehydrogenase (LDH) levels were reported to be associated with a worse outcome in hospitalized patients^20^. Furthermore, low lymphocyte count was also associated with a more severe COVID-19 disease course^21^.

Additionally, as severe cases of COVID-19 suffer from a “cytokine storm“^22,23^ which is an abnormal regulation and excessive release of different pro-inflammatory cytokines as a response to the infection, various cytokines were identified as biomarkers predictive of severe COVID-19. IL-6 is known to play a role as a prognostic marker in COVID 19 since most patients show elevated IL-6 levels during a SARS-CoV-2 infection^23–25^. Several studies have shown high IL-10 levels in severe COVID-19^26,27^. Similarly, IL-17A^28^ and tumor necrosis factor alpha (TNF-α)^25,29^ were reported to be associated with disease severity and progression. As to type-I interferon response, it was reported that hospitalized patients with increased IFN-α levels showed improvement in the COVID-19 disease course^30^. In contrast, high IFN-γ levels in hospitalized cases were shown to be correlated with a worse diagnosis^31^. We had previously reported that the ratio between type-I interferon response and inflammatory cytokines, at the day of hospitalization, shows the highest accuracy for predicting COVID-19 severity and mortality^32^.

However, all the above studies only tested biomarkers predicting severity in patients that were already hospitalized. It would be rather highly beneficial to discover biomarkers for early predictors of severity at the earliest time possible (i.e., as soon as possible after the first positive SARS-CoV-2 diagnosis and within 5-7 DPSO), which are feasible to measure at the point-of-testing. Thus, here we are combining biochemistry, hematological and cytokine biomarkers to predict as early as possible the risk of hospitalization, and the symptomatic versus asymptomatic course of infection.

## Methods

### Patients and study cohorts

The Early-opt-COVID-19 study, a longitudinal study was performed at the University Hospital Augsburg and approved by the local ethics committee of the Technical University of Munich (internal code 799/20 S). Adult study participants were enrolled after written informed consent. Patients were included at the day of the SARS-CoV-2 PCR test at the test center or within 1-5 days after the PCR test through passive recruitment via flyers. The main inclusion criteria were either the presence of COVID-19-related symptoms or contact with a SARS-CoV-2 infected person within the last five days. Exclusion criteria encompassed individuals with chronic virus infections, individuals taking immunosuppressants, and individuals with immune deficiency or pregnancy. Only non-vaccinated and pre-Omicron infected patients were included in the analysis, in order to avoid the effect of a COVID-19 vaccination or infection with less severe variants such as omicron (B.1.1.529). Patients were divided into two main groups, non-hospitalized and hospitalized patients.

Visit 1 at the day of PCR test, for patients recruited at the test center, was available for 36 patients, while the other patients (n=56) started at visit 2 (1-5 days after PCR test), with following visit 3 (2-3 weeks after) and visit 4 (4-5 weeks after). Only patients with their first sample available at DPSO≤11 were included in the analysis, since at later time points there was a significant decline in the various biomarker levels. The patients completed a symptoms questionnaire at each visit to classify the disease course.

From 1.2.2021 until 14.9.2021, n=115 patients were recruited in Augsburg, of which 83 non-hospitalized patients were recruited within the Early-Opt-COVID-19 study, n=9 non-hospitalized patients were included within the COVID-19 Vaccine Consortium (CoVaKo) study^33^ and 23 hospitalized patients were included at the University Hospital Augsburg. In addition, we used results from our previous study for n=70 patients hospitalized at the Pitié-Salpêtrière hospital in Paris^32^. Additionally, n=14 control subjects with negative by SARS-CoV-2 PCR test were enrolled. Only patients with the first visit at most 11 days post symptoms onset (DPSO average 6.2, IQR: 4-9) were included.

Demographic and clinical patient characteristics are summarized in Table 1. Hospitalized patients were divided into three groups according to the oxygen support needed. Patients ventilated with nasal cannula, oxygen mask or without oxygen support were grouped as No-MVS (n=36). Mechanical ventilatory support (MVS, n=43) patients required invasive mechanical ventilation and ECMO patients (n=14) patients required extracorporeal membrane oxygenation^32^. The non-hospitalized patients were divided into 3 symptom groups (Table 1 and Supplementary Figure 1): no symptoms (asymptomatic), mild and few flu-like symptoms (low-symptomatic), and at least 2 out of 5 severe (confusion, shortness of breath, fever, fatigue, diarrhea) symptoms (high-symptomatic), according to the Kings College^16^. Low-symptomatic patients exhibited on average 3.75 symptoms, significantly (p<0.001) lower than 7.6 symptoms in high-symptomatic patients. More details in supplementary methods.

**Table 1:**
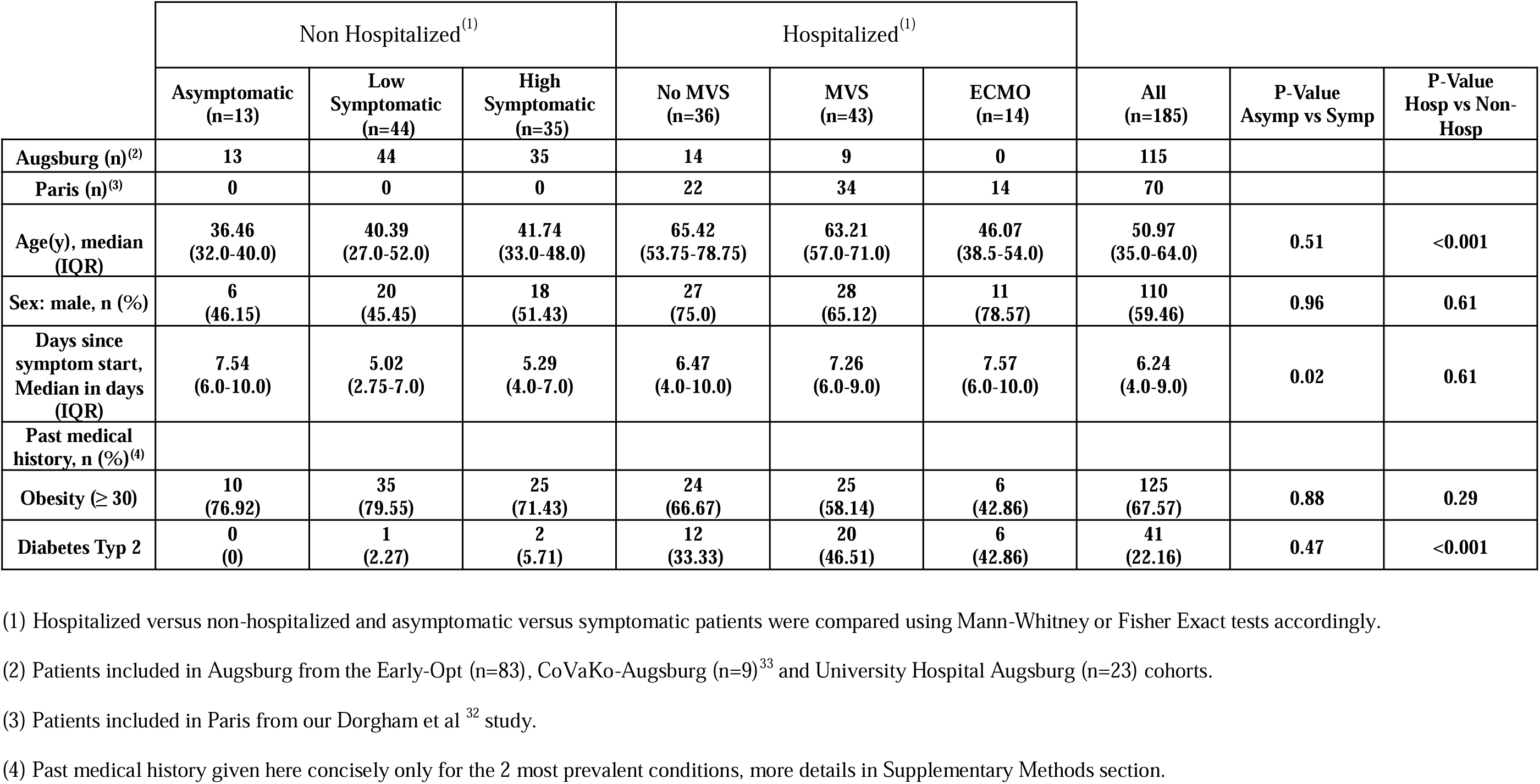
Patient demographics and clinical parameters by Covid-19 symptoms and severity groups.

|  | Non Hospitalized<sup>(1)</sup> |  |  | Hospitalized<sup>(1)</sup> |  |  | All<br>(n=185) | P-Value<br>Asymp vs Symp | P-Value<br>Hosp vs Non-Hosp |
| --- | --- | --- | --- | --- | --- | --- | --- | --- | --- |
|  | Asymptomatic<br>(n=13) | Low<br>Symptomatic<br>(n=44) | High<br>Symptomatic<br>(n=35) | No MVS<br>(n=36) | MVS<br>(n=43) | ECMO<br>(n=14) |  |  |  |
| Augsburg (n)<sup>(2)</sup> | 13 | 44 | 35 | 14 | 9 | 0 | 115 |  |  |
| Paris (n)<sup>(3)</sup> | 0 | 0 | 0 | 22 | 34 | 14 | 70 |  |  |
| Age(y), median<br>(IQR) | 36.46<br>(32.0-40.0) | 40.39<br>(27.0-52.0) | 41.74<br>(33.0-48.0) | 65.42<br>(53.75-78.75) | 63.21<br>(57.0-71.0) | 46.07<br>(38.5-54.0) | 50.97<br>(35.0-64.0) | 0.51 | <0.001 |
| Sex: male, n (%) | 6<br>(46.15) | 20<br>(45.45) | 18<br>(51.43) | 27<br>(75.0) | 28<br>(65.12) | 11<br>(78.57) | 110<br>(59.46) | 0.96 | 0.61 |
| Days since<br>symptom start,<br>Median in days<br>(IQR) | 7.54<br>(6.0-10.0) | 5.02<br>(2.75-7.0) | 5.29<br>(4.0-7.0) | 6.47<br>(4.0-10.0) | 7.26<br>(6.0-9.0) | 7.57<br>(6.0-10.0) | 6.24<br>(4.0-9.0) | 0.02 | 0.61 |
| Past medical<br>history, n (%)<sup>(4)</sup> |  |  |  |  |  |  |  |  |  |
| Obesity (≥ 30) | 10<br>(76.92) | 35<br>(79.55) | 25<br>(71.43) | 24<br>(66.67) | 25<br>(58.14) | 6<br>(42.86) | 125<br>(67.57) | 0.88 | 0.29 |
| Diabetes Typ 2 | 0<br>(0) | 1<br>(2.27) | 2<br>(5.71) | 12<br>(33.33) | 20<br>(46.51) | 6<br>(42.86) | 41<br>(22.16) | 0.47 | <0.001 |

(1) Hospitalized versus non-hospitalized and asymptomatic versus symptomatic patients were compared using Mann-Whitney or Fisher Exact tests accordingly.
(2) Patients included in Augsburg from the Early-Opt (n=83), CoVaKo-Augsburg (n=9)<sup>33</sup> and University Hospital Augsburg (n=23) cohorts.
(3) Patients included in Paris from our Dorgham et al <sup>32</sup> study.
(4) Past medical history given here concisely only for the 2 most prevalent conditions, more details in Supplementary Methods section.

### Biological sampling

On visit 1, only one serum Gel S-Monovette (SARSTEDT) was drawn from the patients. On visits 2-4, four serum Gel S-, a citrate, and an EDTA Monovettes of blood were drawn. On visits 2-4 an oropharyngeal swab was taken and stored in Guanidine Thiocyanate (PrimeStore MTM) medium for RNA stabilization. More details in supplementary methods.

### Biochemistry, hematology, immunoassays and SARS-CoV-2 variant

From two serum Gel S, a citrate and an EDTA Monovette, biochemistry, hematology and differential blood count measurements for: CRP, LDH, creatine kinase (CK), platelet (PLT), ferritin, percent of lymphocytes count and neutrophils count, were performed at the University Hospital Augsburg by standard clinical procedures.

Serum Gel S-Monovette was used to assess cytokines, chemokines and interferons levels. IFN-γ, IL-6, IL-8, IL-10, IL-22, and TNF-α were measured using multiplex planar array immunoassay (CorPlex Human Cytokines 10-Pley Panel on SP-X Platform Quanterix). IFN-α and IL-17A were measured by ultra-sensitive single-plex bead-based Simoa assay (HD-1 Analyser, Quanterix).

SARS-CoV-2 variant detection was performed by cDNA synthesis followed by ARTIC-PCR, Nextera XT (Illumina) library preparation and sequencing on Illumina NextSeq 1000. More details in supplementary methods.

### Bioinformatics and statistical analysis

Descriptive and statistical analyses were performed using R and Python. Batch correction was applied to mitigate batch effect bias. Principal component analysis (PCA) was used to cluster patients and compute principal component weights for each parameter. Predictive value of the various biomarkers was evaluated by the Youden J-index (Informedness, Specificity + Sensitivity -1), since accuracy is to be taken only indicatively due to the ratios between number of patients in the different groups does not represent real world ratios. The threshold used for prediction was selected as the point with the highest J-index on the ROC curve for each biomarker. Longitudinal data was analyzed using piecewise linear regression^34^ and mean negative control data was used as a baseline. Statistical significance between groups was evaluated using the nonparametric Mann-Whitney-U test for continuous variables and the Fisher-Exact test for discrete variables. Correlations were evaluated using the nonparametric Spearman test. A two-sided p-value lower than 0.05 was considered statistically significant. Since 19 biomarkers and 5 biomarker combinations were tested, by the Bonferroni rule a p-value of lower than 0.003 was considered significant after multiple testing correction. More details in supplementary methods.

## Results

### Hospitalization risk prediction

Of all blood biochemistry and hematology biomarkers measured at the first sample per patient (0-11 days post symptoms onset), LDH and CRP showed the most significant (p<0.001) elevation in hospitalized versus non-hospitalized patients (Fig 1A-B), and were the only ones to show accurate prediction of hospitalization risk with a Youden J-index larger than 0.8 (LDH: J-index=91.8%, sensitivity=96.2% and specificity=95.6%; CRP: J-index=87.8%, sensitivity=94.4% and specificity=93.4%; Fig 1E, Table 2). D-dimers, ferritin, creatinine kinase and neutrophils percentage were also significantly elevated, while lymphocytes percentage was significantly lower, in hospitalized patients, but none of those showed a good prediction (Supp Figures 2,4,7).

**Figure 1:**
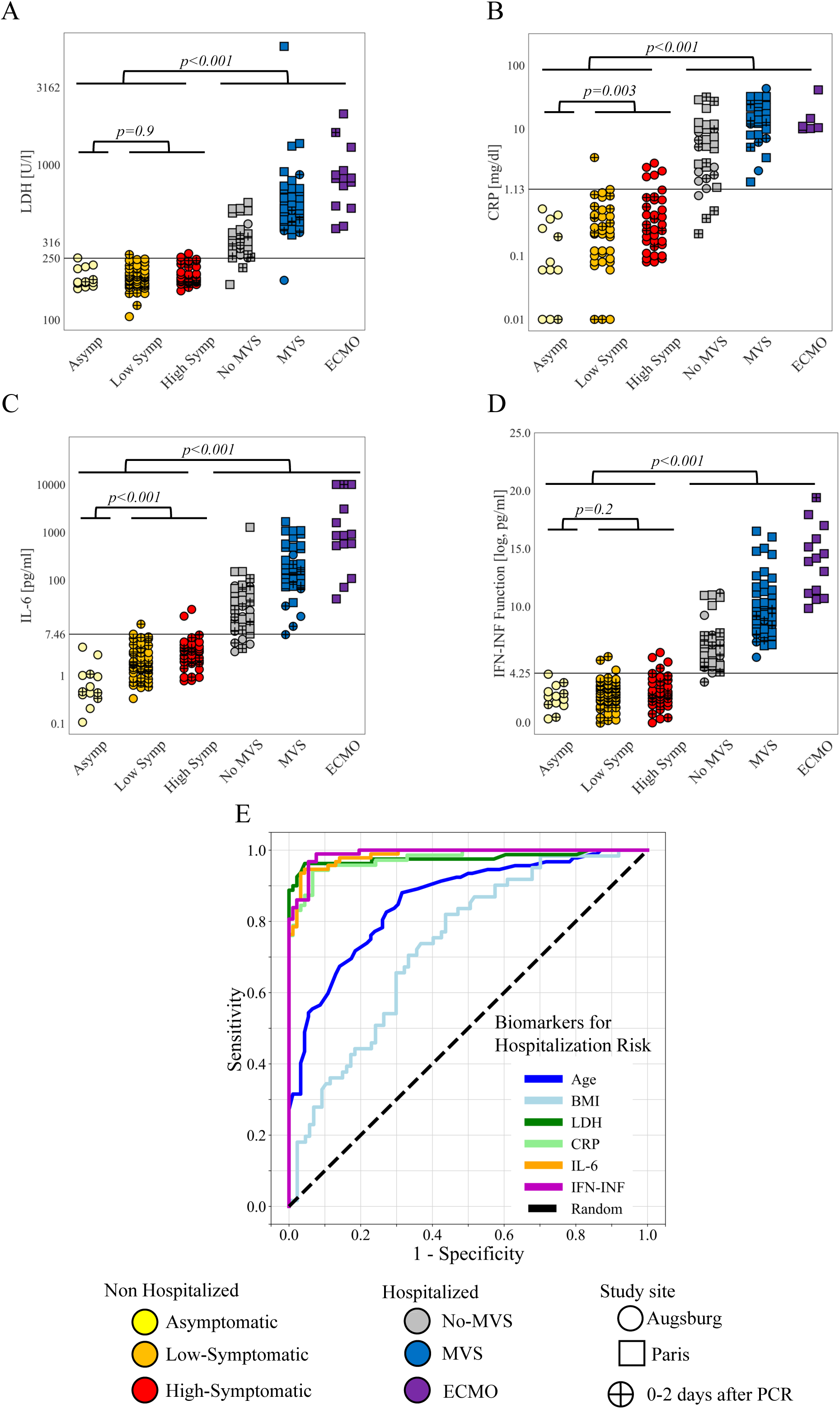
Biochemistry and cytokine biomarkers predictive of COVID-19 hospitalization risk. Early levels of LDH (A), CRP (B) and IL-6 (C), depicted as function of symptoms and severity groups, show significant differentiation between hospitalized and non-hospitalized patients. Also, the ratio between inflammatory cytokine (IL-6, IL-10 and TNF-α) levels compared to IFN-α (IFN-INF function, see Supp Figure 5) is significantly higher in hospitalized patients (D). Receiver Operating Characteristic (ROC) curves for IFN-INF, LDH, IL-6 and CRP show significantly larger AUC for the prediction of hospitalization risk as compared to age and BMI (E). Solid horizontal black lines (A, B, C, D) represent the threshold for each biomarker for which the hospitalization risk prediction has the largest Youden J-index (see Table 2). All other biochemistry and cytokine biomarkers show lower prediction of hospitalization risk (Supp Figures 2,3,4,7). Crossed circles depict patients measured already at the day of the COVID-19 PCR test or within 2 days after. IFN-INF function is defined by: log(2 * IL-6 * IL-10 * TNF-α / IFN-α).

**Figure 2:**
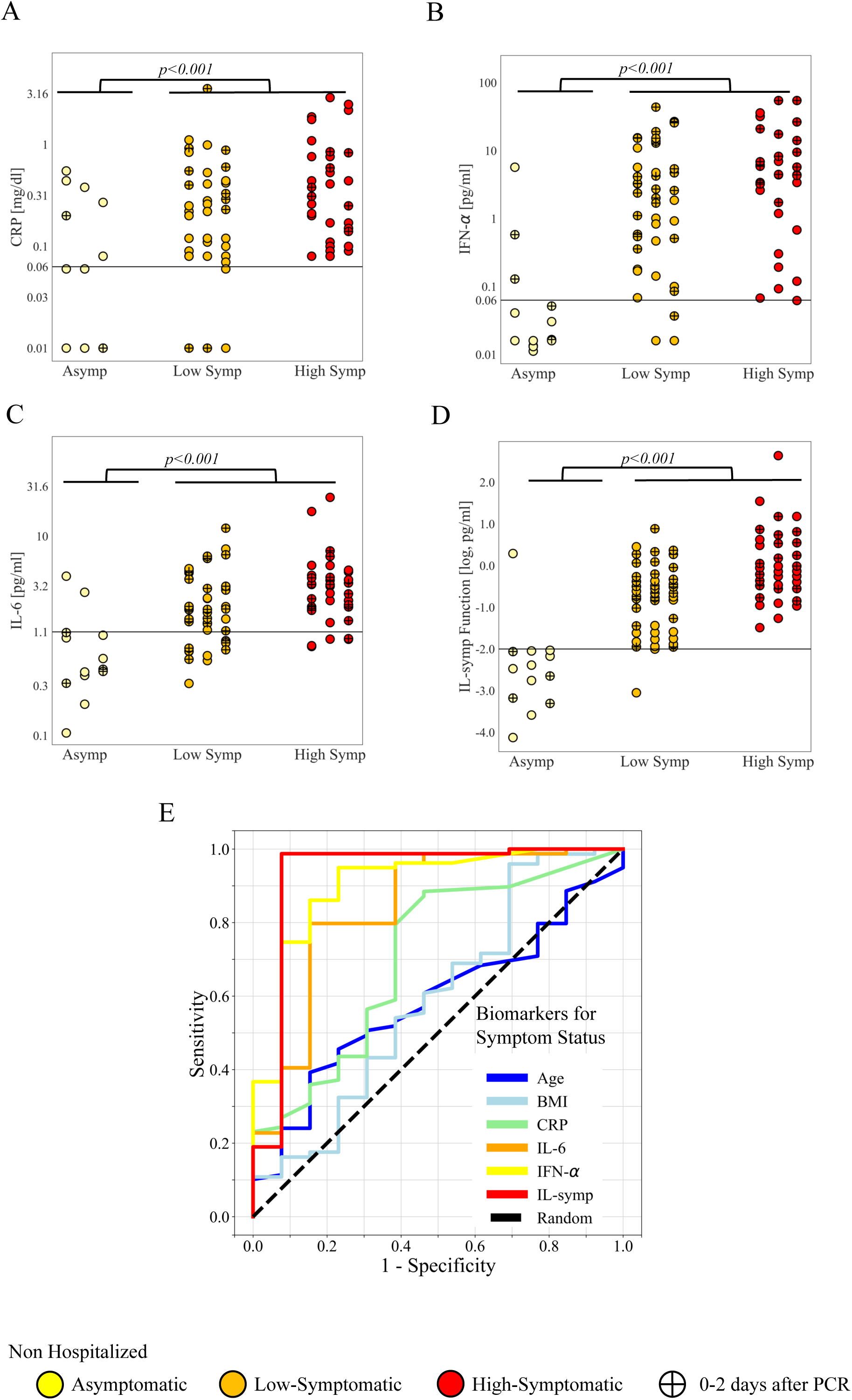
Biochemistry and cytokine biomarkers predictive of COVID-19 symptoms risk. Early levels of CRP (A), IFN-α (B) and IL-6 (C), depicted as function of symptoms groups, show significant differentiation between symptomatic and asymptomatic patients. Also, the product of IL-6, IL-17A, IFN-γ and IFN-α cytokine levels (IL-symp function, see Supp Figure 5) is significantly higher in symptomatic patients (D). Receiver Operating Characteristic (ROC) curves of the prediction for symptoms risk show largest AUC for IL-symp as compared to IFN-α, IL-6, CRP, age and BMI (E). Solid horizontal black lines (A, B, C, D) represent the threshold for each biomarker for which the symptoms risk prediction has the largest Youden J-index (see Table 2). All other biochemistry and cytokine biomarkers show no prediction of symptoms risk (Supp Figures 2,3,4,7). Crossed circles depict patients measured already at the day of the COVID-19 PCR test or within 2 days after. IL-symp function is defined by: -1.6*(log(IL-6)+0.175*log(IFN-γ)) + (log(IL-17A)+0.3*log(IFN-α)).

**Table 2:**
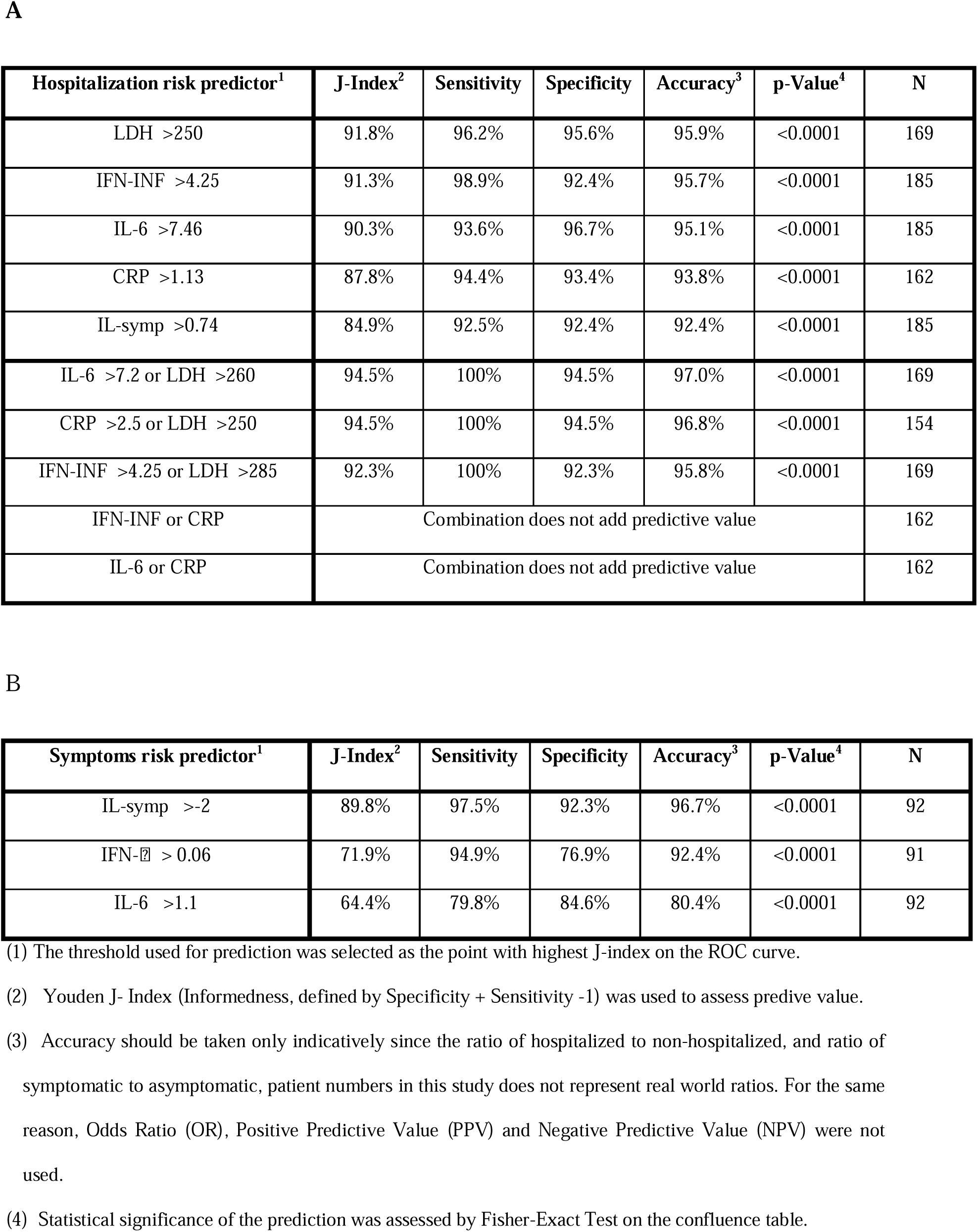
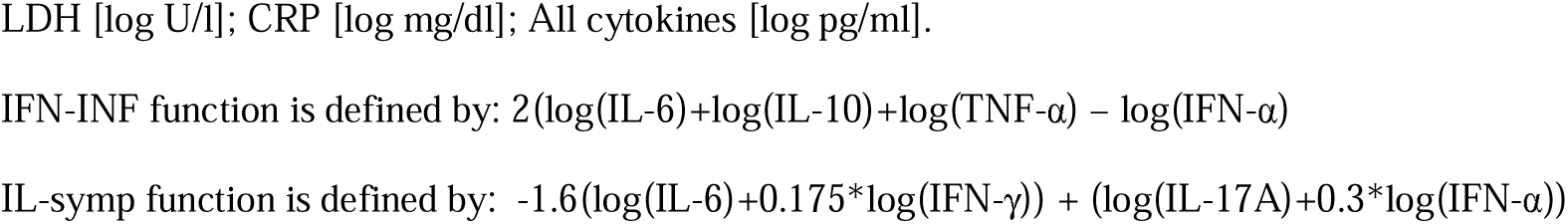
Biomarkers and biomarker combinations allowing early accurate prediction of COVID-19 hospitalization risk (A) or of symptoms risk in non-hospitalized patients (B).

Among the 8 cytokines measured, serum IL-6 level was the most significantly (p<0.001) elevated in hospitalized patients (Fig 1C), and the only cytokine that accurately predicted hospitalization risk (J-index=90.3%, sensitivity=93.6% and specificity=96.7%; Fig 1E and Table 2). IL-8, IL-10, IL-17A, IL-22, TNF-α and IFN-g were also significantly elevated in hospitalized versus non-hospitalized patients, but none of those showed a good prediction of hospitalization risk (Supp Figures 3,4,7). IFN-α rather shows a trend for lower levels in hospitalized patients, especially in ECMO patients, versus symptomatic patients (Supp Figures 3-4).

**Figure 3:**
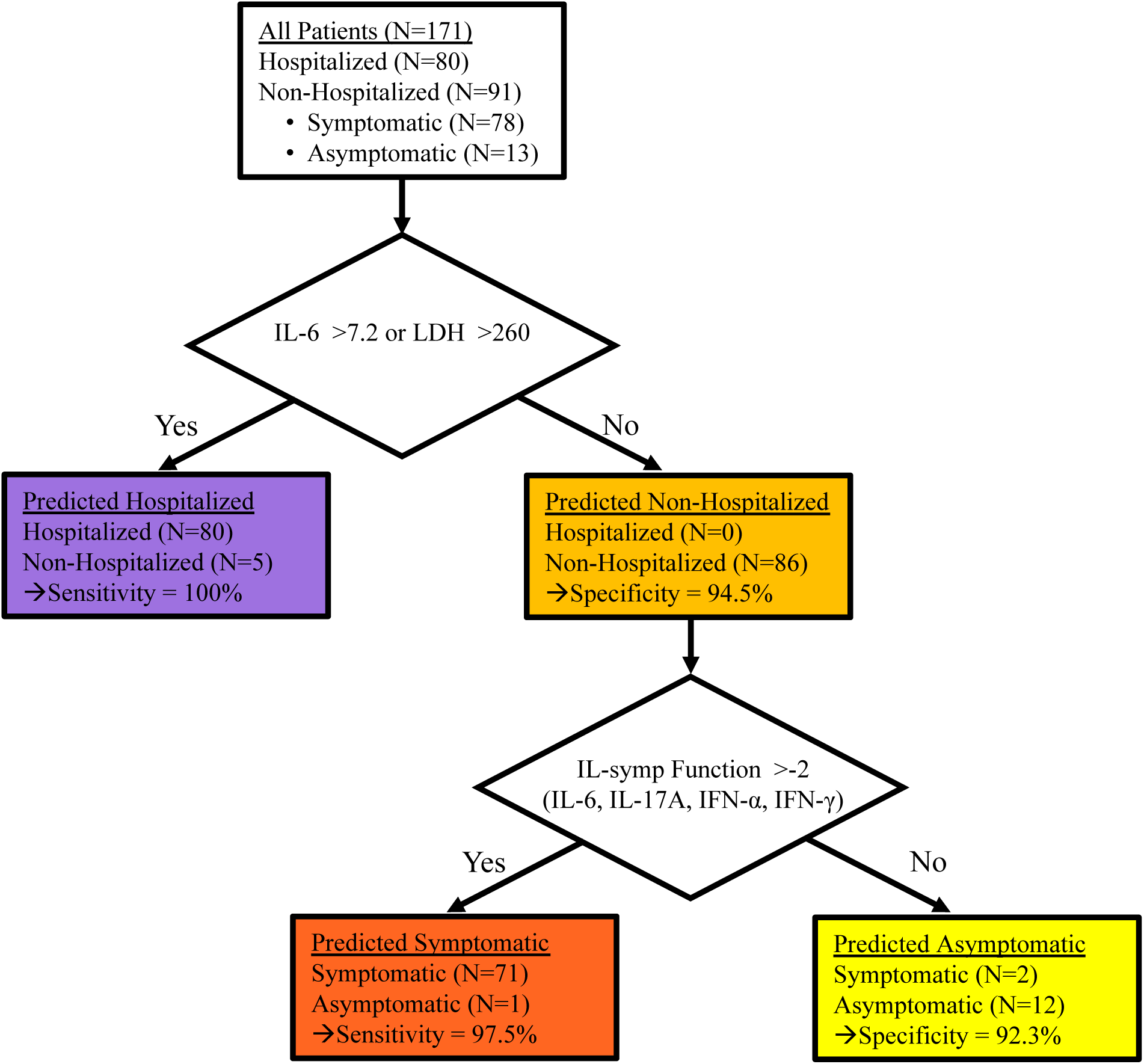
Schematic flowchart of the decision tree for the combined prediction for hospitalization risk and symptoms risk. A flowchart showing the stratification of patients in the study to those predicted (using IL-6 or LDH combination) for hospitalization (with 100% sensitivity) versus non-hospitalization, and the latter further stratified to those predicted (with the IL-symp function) to have symptomatic (sensitivity=97.5%) versus asymptomatic course of infection. Only 169 patients are shown since 26 patients were missing LDH values. IL-symp function is defined by: - 1.6*(log(IL-6)+0.175*log(IFN-γ)) + (log(IL-17A)+0.3*log(IFN-α)).

**Figure 4:**
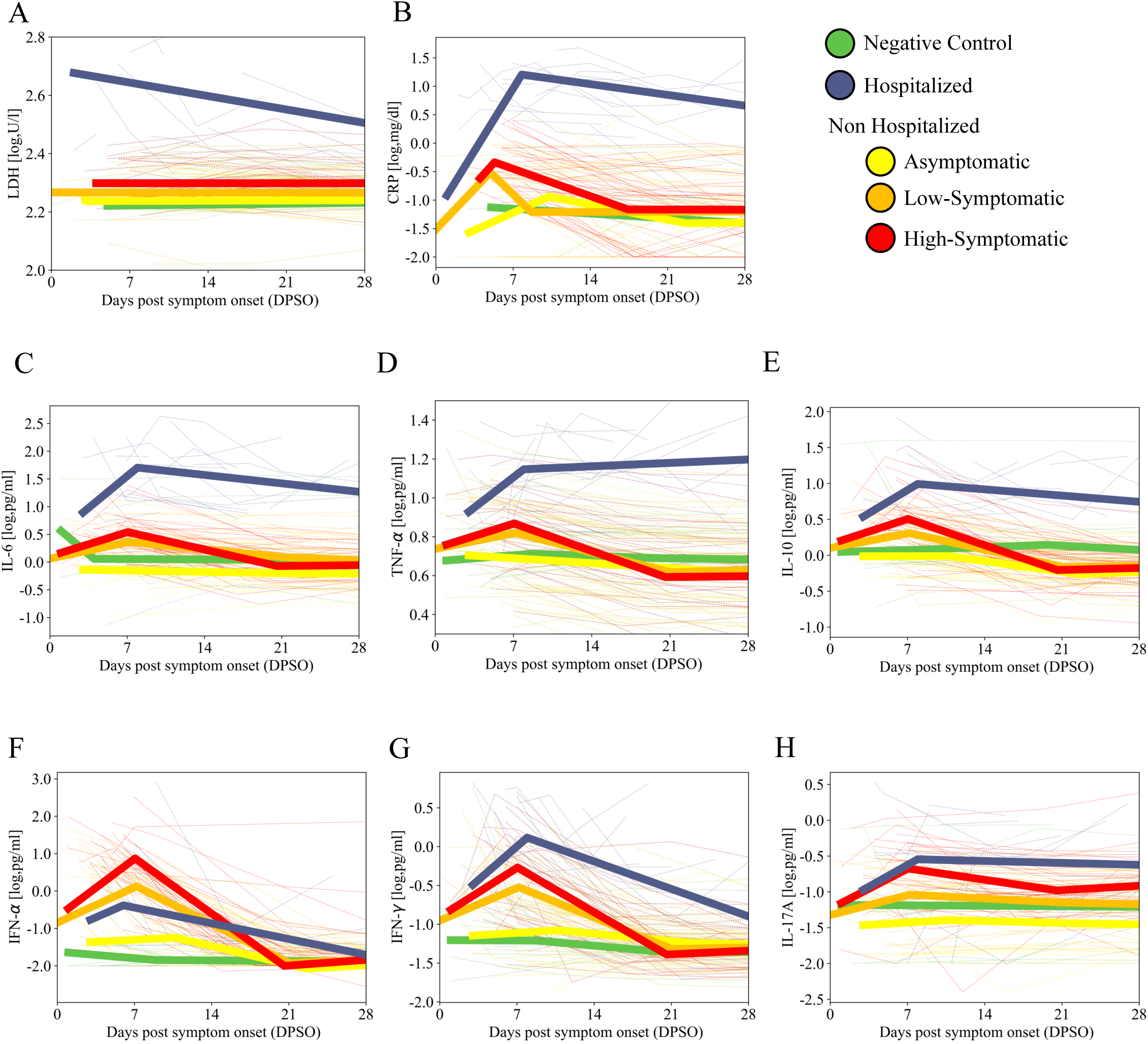
Kinetics of the main cytokine and biochemstry biomarkers stratified by the different severity and symptom groups. Cytokine and biochemstry biomarkers in SARS-CoV-2 positive patients, as well as in the negative control group, were measured 3-4 times over the span of 28 days post symptoms onset (DPSO). Different kinetic profiles for LDH (A), CRP (B), IL-6 (C), TNF-α (D), IL-10 (E), IFN-α (F), IFN-γ (G) and IL-17A (H) are observed in hospitalized, high-symptomatic, low-symptomatic and asymptomatic patients as compared to negative controls, using the average piecewise linear regression of the patients in each group (thick lines). The dotted thin lines in the background represent the kinetics of the single patients.

Next, we used principal component analysis (PCA) for a multidimensional analysis of all the cytokines normalized (Z-scaled) serum levels. Staring with a PCA including all patients and all 8 cytokines, a clear separation was observed between hospitalized and non-hospitalized patients (Supp Figure 5A). Dimension reduction showed a good separation also in PCA with only 4 cytokines (IL-6, IL-10, TNF-α and IFN-α), which was also validated also for the cytokine raw values (Supp Figure 5B). The separatrix line between hospitalized and non-hospitalized patients was then translated to a prediction function of the ratio between the inflammatory cytokines and IFN-α, (termed here the IFN-INF prediction function),

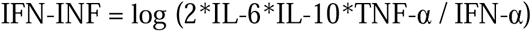

which was significantly (p<0.001) higher in hospitalized patients (Fig 1D) and resulted in accurately predicting hospitalization risk with J-index=91.3% (sensitivity=98.9% and specificity=92.4%; Fig 1E and Table 2). The combination of 3 inflammatory cytokines as a ratio to IFN-α in the IFN-INF function somewhat increases the predictive value compared to IL-6 alone.

ROC analysis comparison of the prediction obtained by each cytokine alone or each of the biochemistry and hematology parameters as well as the IFN-INF combined cytokines prediction function (Fig 1E, Supplementary Figure 7), shows that LDH and the IFN-INF function have the largest area under the curve, although not statistically significantly better than CRP or IL-6 alone. Age and BMI are significantly higher in hospitalized patients but show significantly lower predictive value for hospitalization risk (Supp Figures 7-8).

While LDH, CRP, IL-6, and the IFN-INF function, are all correlated with age and BMI, their predictive value is independent of these factors or of gender and diabetes status (Supp Fig 8-9). Importantly, these 4 predictors are not correlated with, and their predictive value is independent of, the time the sample was taken at the range of 0-11 DPSO or the study site (Supp Figures 8-9). Also, the separation between hospitalized and non-hospitalized patients for these 4 biomarkers is observed over all SARS-CoV-2 variants in the study (Supp Figures 8-9).

Lastly, we investigated if combining the biochemistry and cytokine biomarkers increases the prediction accuracy. The IFN-INF cytokine ratio, IL-6, LDH and CRP are all significantly correlated with each other (Supp Figure 6). Nevertheless, combining (with an OR function) LDH with either IL-6 or the IFN-INF function allows the increase of sensitivity to 100% with only a slight reduction of specificity to 92.3-94.5% (Table 2). Similarly, combining (LDH or CRP) gives sensitivity=100% and specificity=94.5%, although it needs to be noted that CRP data was not available for 12% of patients. All other double or triple biomarker combinations do not add predictive value to the corresponding single biomarker predictions.

### Symptom risk prediction in non-hospitalized patients

Next, we focused on non-hospitalized patients to discover a biomarker predicting symptomatic versus asymptomatic course of infection. CRP, but not LDH or any other biochemistry biomarker, was significantly elevated in symptomatic versus asymptomatic patients (Fig 2A, Supp Figures 2,4). Platelet count was significantly lower in symptomatic patients (Supp Fig 2,4). None of the biochemistry, hematology or differential blood count biomarkers was predictive for symptoms risk in non-hospitalized patients (Supp Fig 7D).

Among the cytokines, IFN-α, IFN-γ, IL-6, IL-10, IL17-A and IL-22 were significantly elevated in symptomatic versus asymptomatic patients (Fig 2B-C, Supp Fig 3-4). However, only a modest (J-index= 64-72%) predictive value for symptomatic risk was found only for IFN-α and IL-6, but for none of the other cytokine nor for the IFN-INF cytokine ratio (Fig 2E, Table 2, Supp Fig 7C).

Therefore, we investigated whether another combination of cytokines could predict the symptomatic risk. PCA performed for only non-hospitalized (asymptomatic versus symptomatic) patients with all 8 cytokines shows some separation between these groups (Supp Figure 5C). Using the same methods and dimension reduction as for the IFN-INF prediction, we found that IFN-α, IL-17A, IL-6, and IFN-γ are the most important cytokines for separation between asymptomatic patients and symptomatic patients. Translation into raw values of these cytokines shows clear separation between asymptomatic and symptomatic ((Supp Figure 5D), and using the PC factors and the separatrix function parameters we obtained the IL-symp prediction function

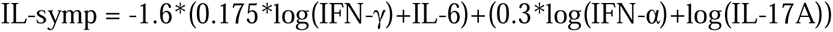

being significantly (p<0.0001) elevated in symptomatic patients (Fig 2D). The symptomatic risk is accurately (J-index= 89.8%) predicted by the IL-symp cytokine function with a sensitivity of 97.5% and a specificity of 92.3% (Fig 2E, Table 2). IL-symp prediction function is also predictive of hospitalization risk but less accurately than LDH, IFN-INF, IL-6 or CRP (Supp Fig 7).

While IL-symp is correlated with age and BMI, its predictive value is independent of these factors or of gender and diabetes status (Supp Fig 10). IL-symp value is somewhat decreased after 7 DPSO, but its predictive value is independent of the time the sample was taken, within the range of 0-11 DPSO, or of the SARS-CoV-2 variant (Supp Fig 10). CRP, LDH or other biochemistry parameters are not strongly correlated with IL-symp, and the combination of IL-symp (or any single cytokine) with the biochemistry biomarkers does not improve its predictive value.

### Decision tree for combined prediction of hospitalization risk and symptomatic status

Thus, by measuring only LDH and 4 cytokines (IL-6, IL-17A, IFN-α and IFN-γ) it is possible to obtain a combined decision tree (Fig 3), predicting in our study hospitalized patients with sensitivity of 100% (using the IL-6 and LDH combination) and among those predicted not to be hospitalized a prediction of symptomatic patients (using IL-symp) with sensitivity of 97.5%.

### Longitudinal profiles of cytokine and biochemistry biomarkers

The longitudinal measurements starting as early as the day of (max 5, median 4.6, days after) the PCR test and up to 5 weeks DPSO, reveal differences in the kinetic profiles of different biomarkers between hospitalized as well as between non-hospitalized and symptomatic and asymptomatic patients. Using piecewise linear regression for each group of patients (except for ECMO patients that had no longitudinal data) the average biomarker kinetics profile per severity or symptom group is given in Figure 4. LDH, CRP, IL-6, IL-10 and TNF-α are elevated in hospitalized in comparison to non-hospitalized patients already at 0-7 DPSO, and stay significantly elevated up to 28 DPSO, with no significant difference between the low- and high-symptomatic patients. On the other hand, IFN-α, IFN-γ and IL-17A levels show an increase with the degree of symptoms, but no significant difference between hospitalized patients and high-symptomatic non-hospitalized patients

## Discussion

Our findings indicate that it is possible to predict COVID-19 disease progression with biomarkers feasibly measurable at point-of-testing already as early as 0-7 days post symptoms onset and even at the time of the first positive diagnosis of SARS-CoV-2 infection. Most importantly, we have shown that highly accurate (J-index >90%) early prediction of COVID-19 hospitalization risk is possible either by high LDH levels, or high IL-6 levels, or by the ratio between IFN-α and the inflammatory cytokines IL-6, IL-10 and TNF-α, termed here the IFN-INF predictor function. Moreover, combining the LDH with IL-6 biomarkers enables a predictor with sensitivity of 100%. It should be noted that all above predictors are not statistically significantly different in their predictive values. While all above biomarkers have been previously shown to predict COVID-19 severity in hospitalized patients^19,20,23–31^,our results now show that they are also good early predictors for hospitalization risk.

Furthermore, we have shown that a combination of the IFN-α, IFN-γ, IL-6 and IL-17 cytokines, termed here the IL-symp prediction function, is an early and accurate (J-index=89.8%) predictor of symptomatic versus asymptomatic course of infection. Interestingly, neither any single cytokine alone, nor the IFN-INF cytokine combination, nor any of the other biochemistry or hematological parameters, were a good predictors for symptoms risk. While the prediction of symptomatic status is clinically less important, still it could be used to guide personalized treatment with SARS-CoV-2 anti-viral therapy, as these are proven in clinical trials to reduce the associated quality-of-life impairment and public-health burden in mild-moderate patients^3–7^.

For a large fraction (39.1%) of the non-hospitalized patients the above predictions are based on samples taken already at the day of the first diagnosis of a positive SARS-CoV-2 at the test center, but none of the patients recruited at the test centers were finally hospitalized. Nevertheless, we show that the above predictions of both hospitalization and symptoms risk are independent of the time of sampling.

We also found that the risk of hospitalization and the risk of symptomatic status are associated with older age and higher BMI, similar to previous studies showing these factors associated with COVID-19 severity^17,18^. However, we show that the predictors we have identified are independent of such factors as age, gender, BMI or diabetes, probably since these are already factorized into the early levels of the biomarkers in response to SARS-CoV-2 infection.

Our results also shed further light on the role of the different cytokines in response to SARS-Cov-2 infection. While high levels of the inflammatory cytokines (e.g. IL-6, IL-8 and TNF-α), and also interestingly IL-10, are associated with both the risk for hospitalization and higher risk for symptomatic course of infection, conversely IFN-α, IFN-γ and IL-17 are less associated with hospitalization risk but rather with the appearance of symptoms. In fact, an early high ratio of IFN-α compared to inflammatory cytokines seems to decrease the risk of hospitalization, similarly to our previous findings about COVID-19 severity in hospitalized patients^32^. It is important to note that since we aimed at finding the minimum number of cytokines that allow a good prediction, some cytokines (e.g. IL-8) were not included in our prediction function since they do not add significant predictive value, probably because of high inter-correlations, but still are associated with risk of hospitalization or of symptomatic disease. Also, we found that the IFN-INF cytokine combination function is positively correlated with LDH and CRP, indicating a biological link between the different processes that these are biomarkers for. Interestingly, there is no strong correlation between the IL-symp function, predictive of symptoms’ risk, with biochemistry or hematological biomarkers or lymphocyte counts.

Our longitudinal results show that the kinetics of the cytokines IL-6, TNF-α and IL-10 differ in hospitalized patients, showing higher expression at an early timeframe (already around 0-7 days post symptoms onset) and staying high even 4 weeks after. While it was shown that inflammatory cytokines are high in severe COVID-19 patients^23–27^, we show here that in some patients they are already high very early on, thus indicating that if immune modulatory treatment is to be successful it should be initiated early personalized according to the cytokine profile^11^. As expected, we see also an early rise in IFN-α and IFN-γ, but these are not as significantly different between hospitalized and symptomatic patients and they decline within 2-3 weeks also in hospitalized patients.

The patients analyzed here were all non-vaccinated and recruited prior to the omicron variant appearance. While most of the hospitalized patients in this study were infected with the wild-type virus and the non-hospitalized patients infected with other variants (mostly alpha as well as beta, eta, iota and delta), we also have some hospitalized patients infected with other variants and in general our predictors were not affected by the different variants. Nevertheless, because of the design of our study, we had a bias towards a larger number of symptomatic patients as compared to non-symptomatic patients. Moreover, since none of the patients recruited at the test centers were finally hospitalized, hospitalized patients were recruited at the hospital rather than at the test center, with a larger than realistic ratio of hospitalized to non-hospitalized patients. Due to these imbalances we test our predictors only using sensitivity and specificity, and their combined Youden J-index (Informedness), rather than using accuracy. Considering the above limitations, a larger sequential recruitment study is needed to validate our findings.

Based on our results, we suggest a combined predictive pipeline for early prediction of patients that will become asymptomatic versus symptomatic versus hospitalized. As early as possible after a positive SARS-CoV-2 test, patients should get blood drawn and LDH, IL-6, IL-17A, IFN-a and IFN-g should be measured, which could be performed at point of testing by a multiplex ELISA or other methods. Using these biomarkers the hospitalization risk could be evaluated by LDH combined with IL-6, and the symptomatic risk could be evaluated by the IL-symp function, therefore allowing for personalized treatment with either anti-viral therapy or with cytokine-inhibitor treatment, which can be also guided by the cytokine levels. With these two populations predicted, personalized therapy approaches can be targeted to the patients with high risk in regards to hospitalization and a symptomatic course.

## Data Availability

All data produced in the present study are available upon reasonable request to the authors.

## Source of funding

This study was funded by the Bavarian State Ministry for Science and Art Program for the funding of Corona research (Early-Opt-COVID19project and research networks FOR-COVID and Bay-VOC), by grants from the Helmholtz Association’s Initiative and Networking Fund (KA1-Co-02 “COVIPA” to UP; KA1-Co-06“CORAERO” to CH, MG and AUN), by the European Commission FET Open Grant VIROFIGHT (grant no. 899619), and by the Sanddorf foundation. Work in the Gorochov laboratory was supported by Institut National de la Santé et de la Recherche Médicale (INSERM), Sorbonne Université, Fondation pour la Recherche Médicale (FRM), Paris, France, program ‘‘Investissement d’Avenir’’ launched bythe French Government and implemented by the Agence Nationale de la Recherche(ANR) (programme COFIFERON ANR-21-RHUS-08), by EU Horizon HLTH-2021-DISEASE-04UNDINE project, by Fondation pour la Recherche Médicale, Paris, France (programme Equipe FRM 2022) and by the Département Médico-Universitairede Biologie et Génomique Médicales (DMU BioGen), APHP, Paris, France.

## Acknowledgments

We thank the patients for their participation in the study. We thank the PerForM-REACT Project (funded by the Free State of Bavaria and the European Regional Development Fund - ERDF) for use of devices and consumables. The French-German collaboration was additionally supported by the BayFrance foundation.

## Disclosure statement

UP received personal fees from Abbott, Abbvie, Arbutus, Gilead, GSK, Leukocare, J&J, Roche, MSD, Sanofi, Sobi and Vaccitech. UP is a co-founder and share-holder of SCG Cell Therapy. All other authors declare no conflict of interest. The study was independently designed, run, analyzed and summarized by the authors with no involvement from the funding agencies. The manuscript was written by the authors with no involvement of the funding agencies. The funding agencies did not pay for or were involved in any way of writing the manuscript.

## Abbreviations

AUC: Area under the curve
BMI: Body Mass Index
CI: Confidence Interval
CK: Creatin Kinase
CoVaKo: Corona-Vakzin-Konsortium
COVID: Coronavirus Disease
CRP: C-reactive protein
DPSO: Days Post Symptom Onset
ECMO: Extracorporeal membaren oxygenation
EDTA: ethylene diamine tetraacetic acid
EOC: Early Opt Covid 19 Study
IFN: Interferon
IL: Interleukin
IQR: Interquantile Range
J-Index: Youden Index
LDH: Lactate Dehydrogenase
MVS: Mechanical Ventilation
PCA: Principal Component Analysis
PCR: Polymerace chain reaction
PLT: Platelet
ROC: Receiving Operator Curve
SARS-CoV: severe-acute-respiratory-syndrome-related coronavirus
TNF: Tumor necrosis factor
WBC: White Blood Cell

## Supplementary figure captions

**Supplementary Figure 1:**
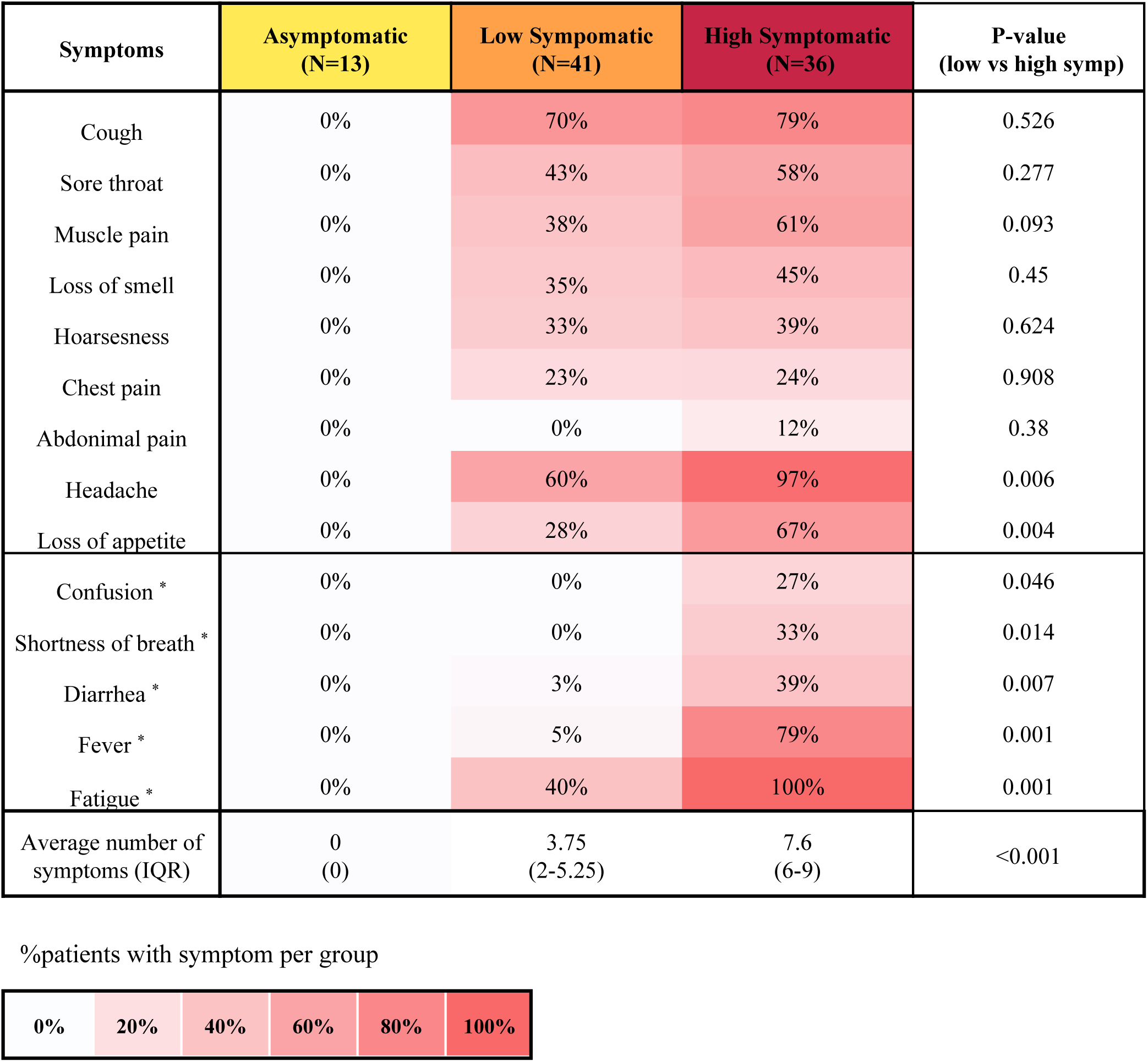
Symptom distribution in different non-hospitalized symptom groups. The distribution of 14 different COVID-19 related symptoms across asymptomatic, low symptomatic and high symptomatic patients in the non-hospitalized cohort. High symptomatic was defined as having at least 2 out of 5 of the symptoms most linked to risk of hospitalization (marked by *). Significant differences are observed between low and high-symptomatic patients in the frequency of several symptoms as well as for the number of symptoms a patient exhibits.

**Supplementary Figure 2:**
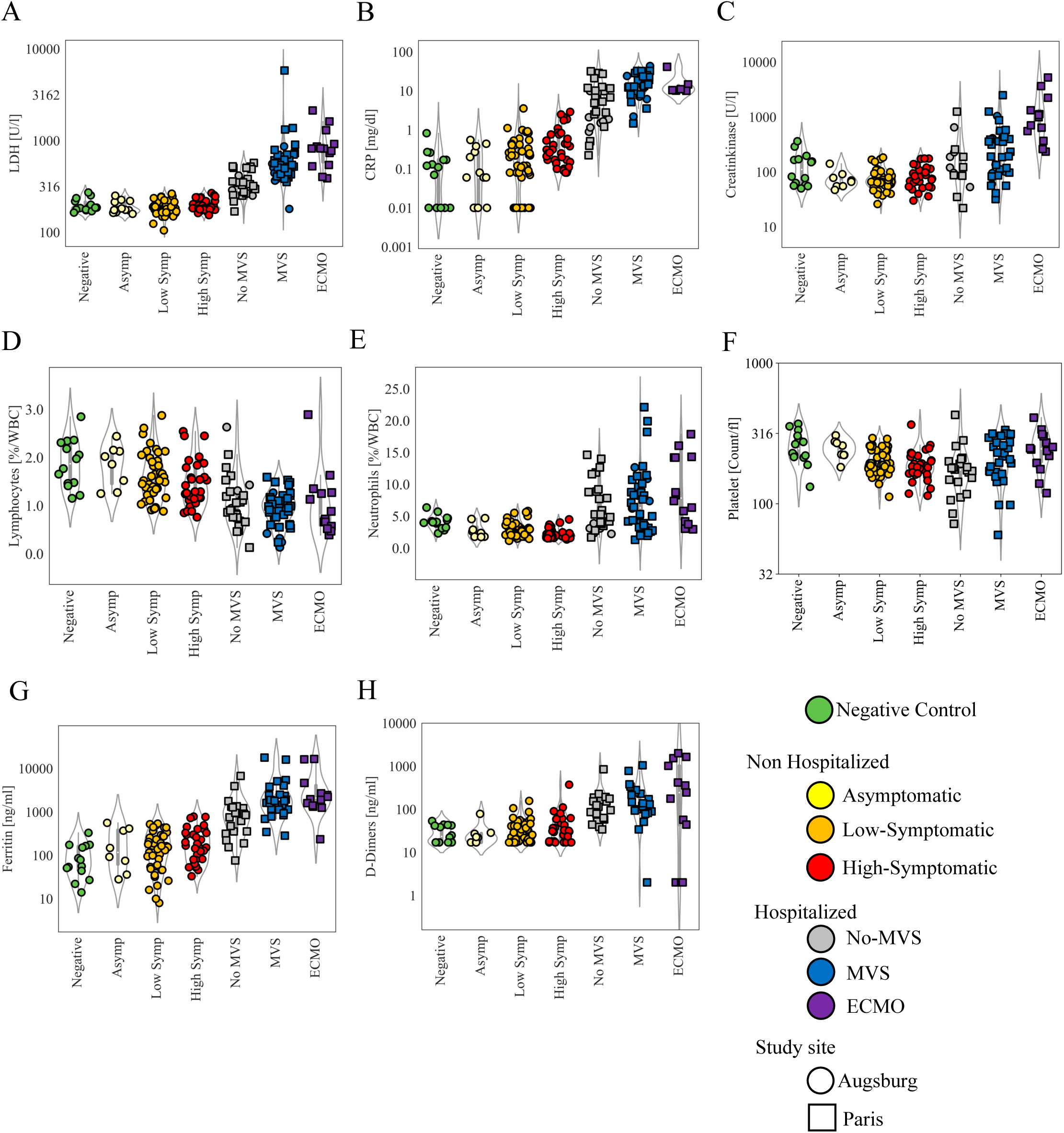
Biochemistry biomarker levels across non-hospitalized symptoms groups and hospitalized severity classes as well as negative control group. The single biomarkers LDH (A), CRP(B), Ceratinkinase (C) , Lymphocytes (D), Neutrophiles (E), Platelets (F), Ferritin (G) and D-dimers (H) show significantly different levels between the severity classes and symptom groups. A wide range of levels for the single markers are also found within the negative patient control group. Circles mark patients from the Augsburg study site and squares for Paris.

**Supplementary Figure 3:**
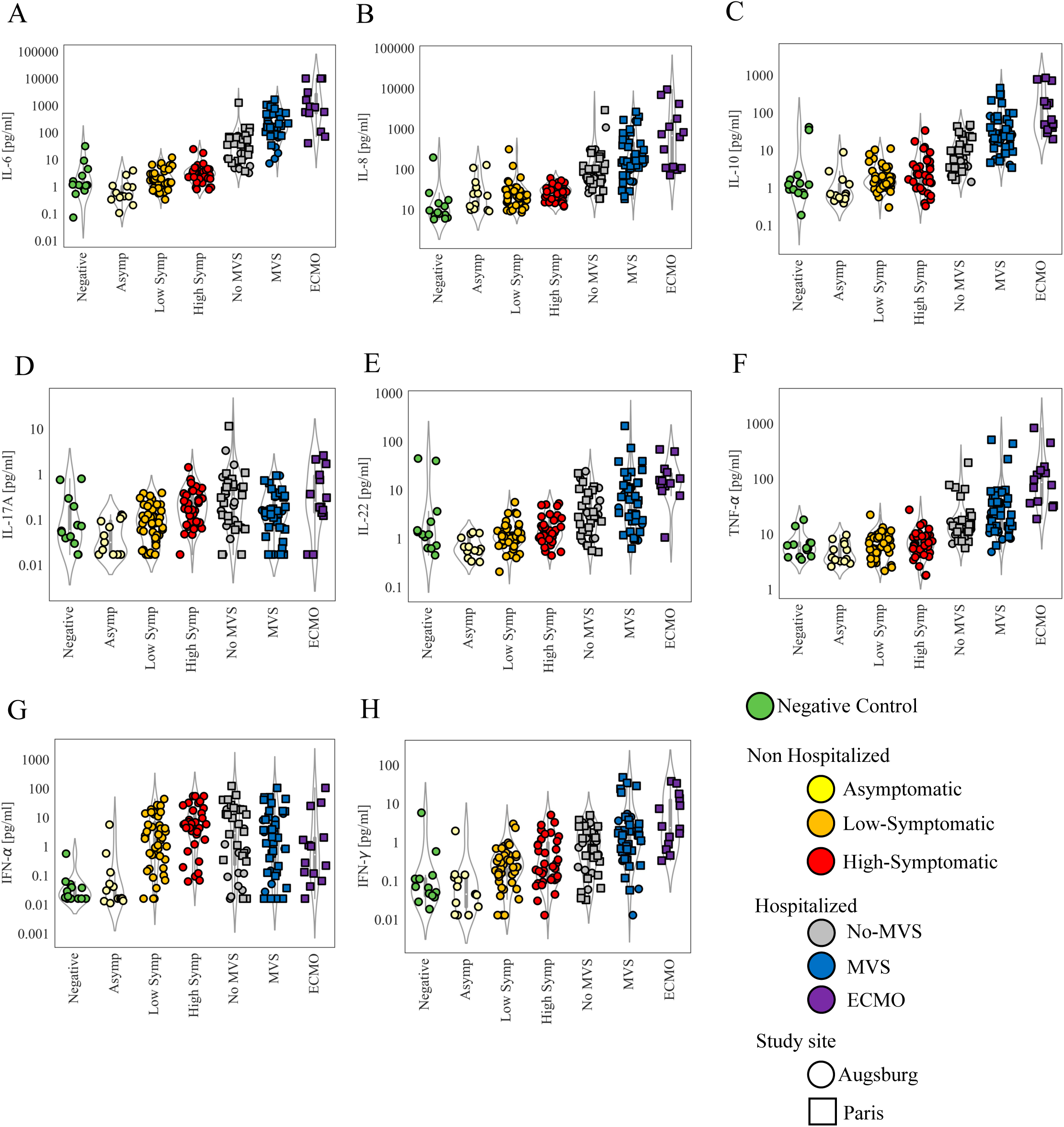
Single cytokine levels across non-hospitalized symptoms groups and hospitalized severity classes as well as negative control group. The levels of single cytokines IL-6 (A), IL-8 (B), IL-10 (C), IL-17A (D), IL-22 (E), TNF-α (F), IFN-α (G) and IFN-γ (H) show significantly different levels between the severity classes and symptom groups. A wide range of levels for the single markers are also found within the negative patient control group. Circles mark patients from the Augsburg study site and squares for Paris.

**Supplementary Figure 4:**
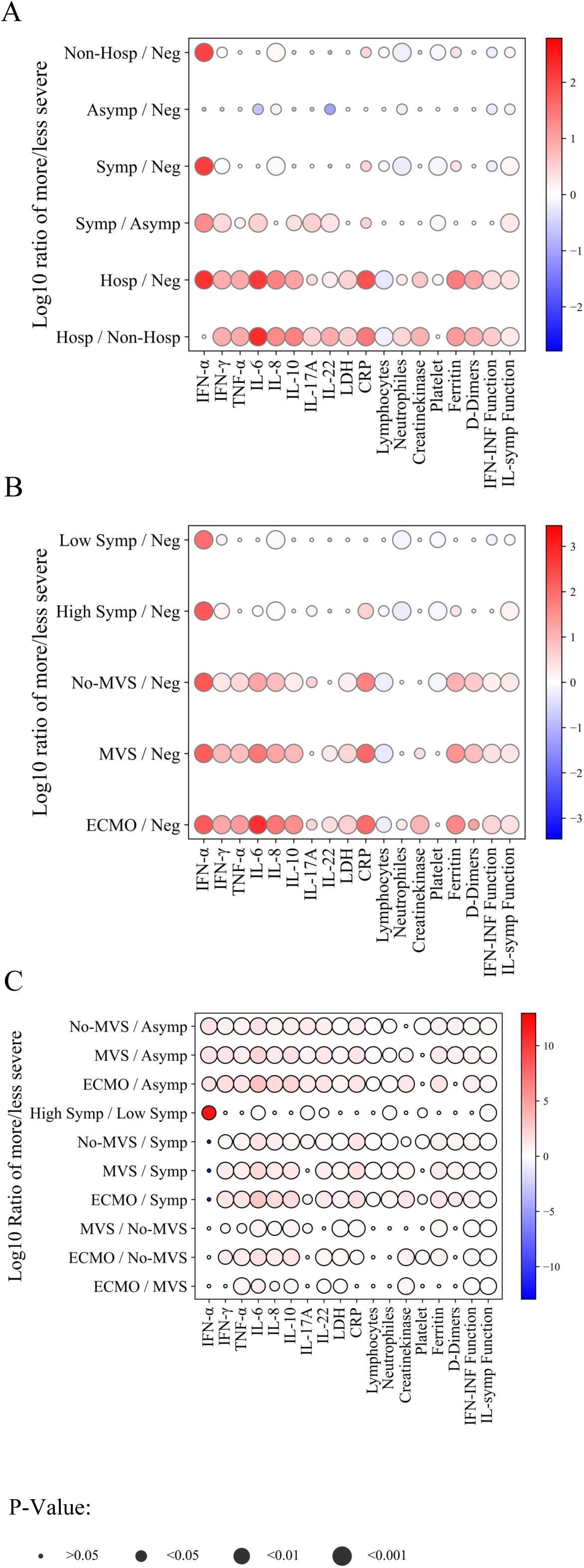
Differences between the levels of cytokine and biochemistry biomarkers across non-hospitalized symptoms groups and hospitalized severity classes as well as negative control group. The difference between the groups is measured by the log2 ratio of more/less severe. The bubble plot depicts the ratio magnitide (bubble color scale) and P-value (bubble size) of the difference between the severity and symptom groups. The order for the groups is asymptomatic, low-symptomatic, high-symptomatic, No-MVS, MVS and ECMO. (A) The differences between the main groups: negative, asymptomatic, symptomatic, non-hospitalized, and hospitalized. (B) The differences between all groups as compated to the negative control groups. (C) Comparison between all sub-groups.

**Supplementary Figure 5:**
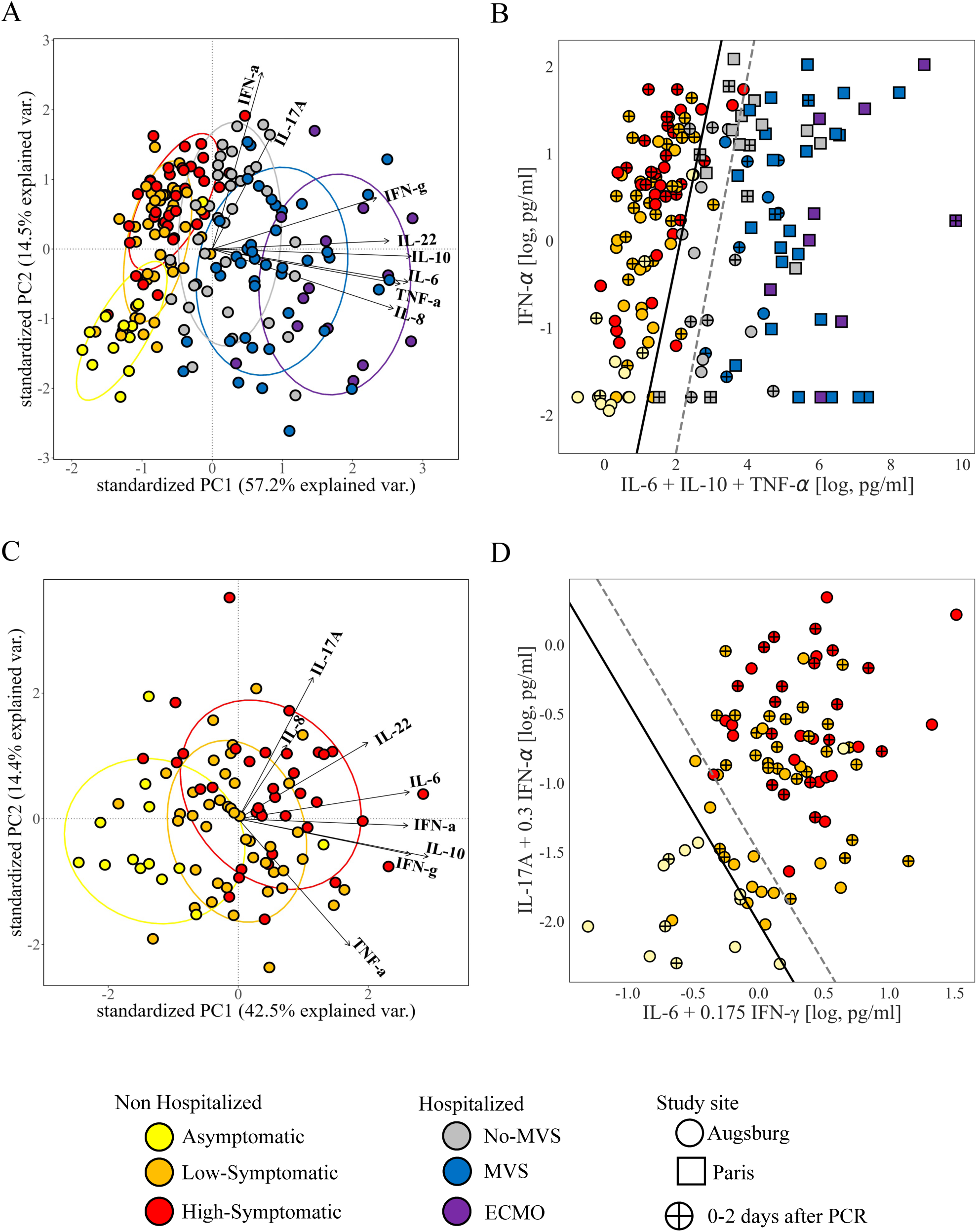
Derivation of the multi-dimensional cytokine combination prediction functions for hospitalizaion risk and symptoms status. PCA of the eight measured cytokines (A) shows differentiation between hospitalized and non-hospitalized patients. After a dimension reduction to determine the minimum number of most predictive cytokines, the ratio between the raw levels of inflammatory cytokine (IL-6, IL-10 and TNF-α) compared to IFN-α (B) shows a clear separation of the 2 groups (solid line), giving rise to the IFN-INF prediction function showing significant prediction of hospitalization risk (Figure 1 and Table 2). A grey zone (between the solid and dashed lines), is observable for IFN-INF function with only No-MVS and high-symptomatic patients (B). Next, PCA of the eight measured cytokines in only non-hospitalized patients (C), shows separation between asymptomatic and symptomatic patients. After a dimension reduction, the product of IL-6, IL-17A, IFN-γ and IFN-α raw levels (D) shows a clear separation of the two groups (solid line), giving rise to the IL-symp prediction function, allowing accurate and significant prediction of symptoms risk (Figure 2 and Table 2). A grey zone (between the solid and dashed lines), is observable for IL-symp function with only low-symptomatic patients (D). Crossed circles depict patients measured already at the day of the COVID-19 PCR test or within 2 days after. PCA ellipses (D) represent 69% of patient distribution in each group. IFN-INF prediction function is defined by: log(2*IL-6*IL-10*TNF-α/IFN-α). IL-symp function is defined by: -1.6*(log(IL-6)+0.175*log(IFN-γ)) + (log(IL-17A)+0.3*log(IFN-α)).

**Supplementary Figure 6:**
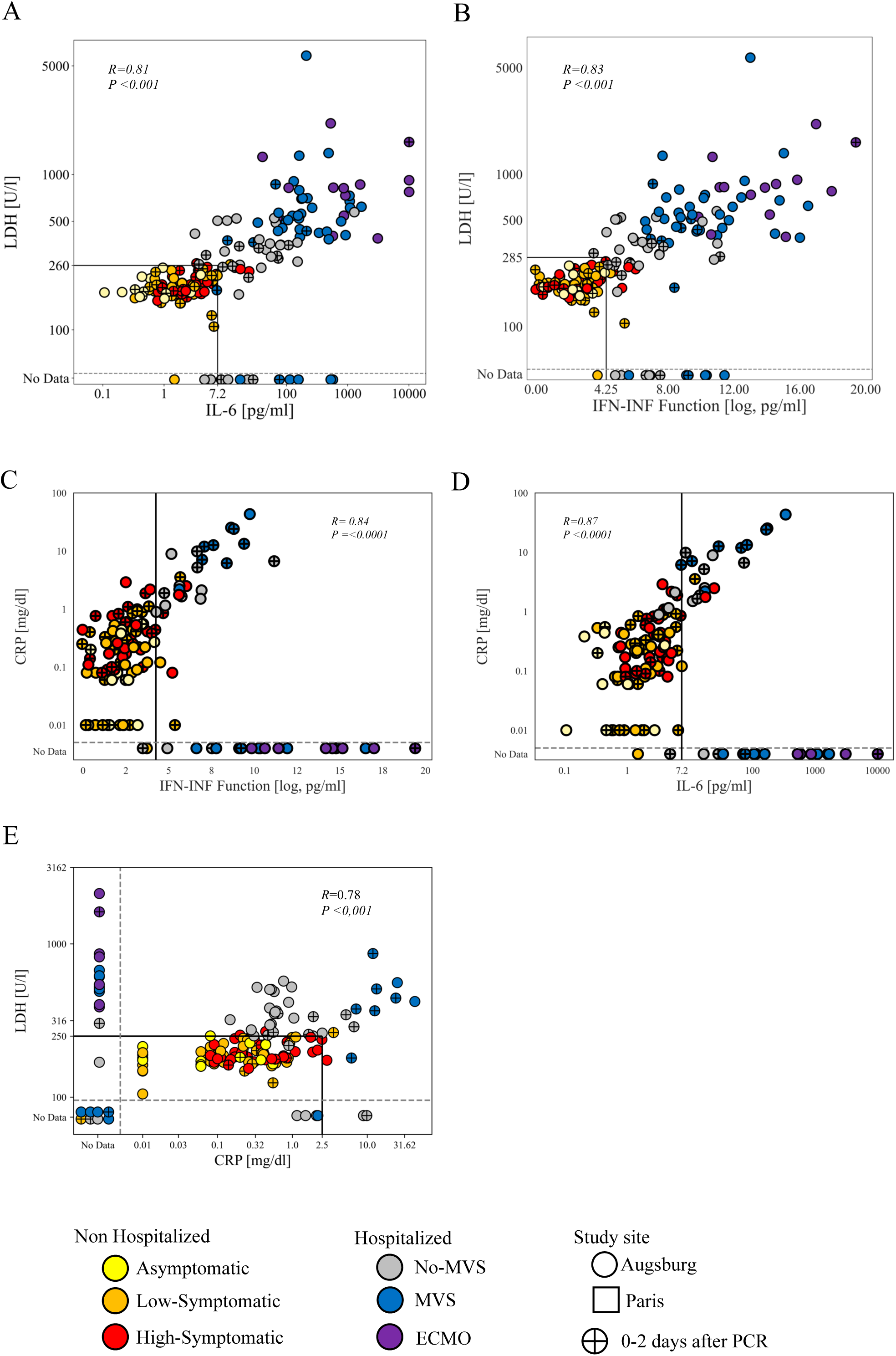
Prediction of hospitalization risk by combining biochemistry and cytokine biomarkers. The combinations of LDH with IL-6 (A) or with the IFN-INF function (B) show a clear separation of hospitalized versus non-hospitalized patients. Consequently, combing the conditions that (LDH>260 or IL-6>7.2, out of square in A) or by (LDH>285 or IFN-INF>4.25, out of square in B) allow for high sensitivity prediction of hospitalization risk. CRP, on the other hand, does not have additive predictive value for IL-6 (C) or IFN-INF function (D). A number of patients are missing LDH and/or CRP data (marked No Data below or to the left of the dotted lines accordingly).

**Supplementary Figure 7:**
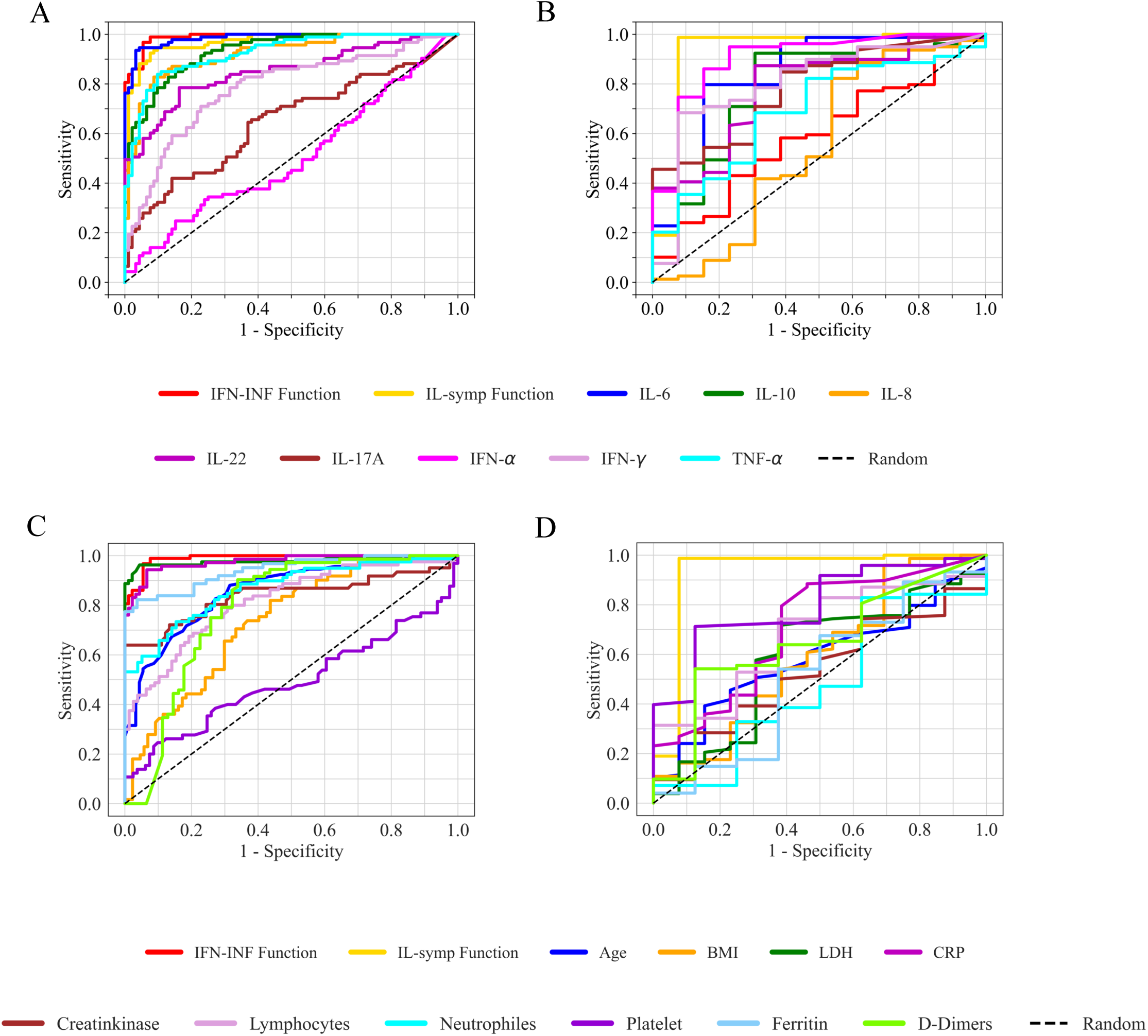
Comparison of the predictive values for hospitalization risk and for symptomatic risk for all measured biochemistry, hematology and cytokine biomarkers. Receiver Operating Characteristic (ROC) curves for the eight measured different single cytokines, as well as the IFN-INF and IL-IEM cytokine combination functions, as predictors for hospitalization risk (A) and for symptomatic risk (B). ROC curves for all biochemistry biomarkers, as well as age and BMI, as predictors for hospitalization risk (C) and for symptomatic risk (D). IFN-INF function is defined by: log(2*IL-6*IL-10*TNF-α/IFN-α). IL-symp function is defined by: -1.6*(log(IL-6)+0.175*log(IFN-γ)) + (log(IL-17A)+0.3*log(IFN-α)).

**Supplementary Figure 8:**
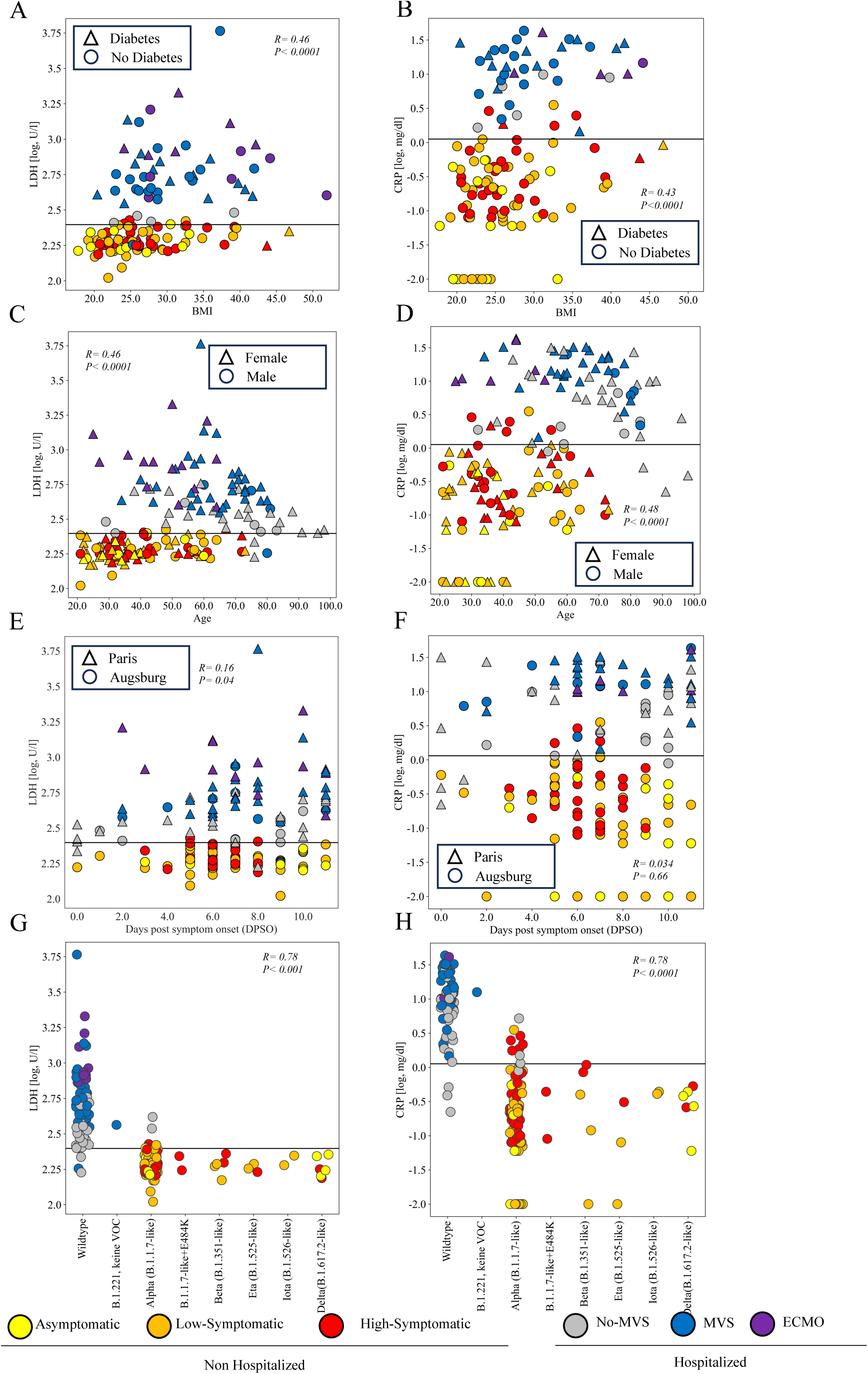
Independence of LDH and CRP as predictors for hospitalization risk as function of different known risk factors for COVID-19 severity. Scatterplots show the independence from other factors of the prediction of hospitalization risk by LDH (A, C, E and G) and CRP (B, D, F and H). Solid horizontal black lines represent the threshold for each biomarker for which the symptoms risk prediction has the largest Youden J-index (see Table 2). Although both LDH and CRP are significntly correlated with BMI (A,B) and age (C,D), prediction by LDH and CRP is independent of BMI (A,B), having diabetes (A,B), age (C,D), or gender (C,D). Importantly, no differences in the prediction power by LDH or CRP are observed as function of time from symptom onset ranging 0-11 DPSO (E,F), or between the study sites (E, F). Lastly, the prediction of hospitalization risk by LDH and CRP held for differrent SARS-CoV-2 variants (G,H).

**Supplementary Figure 9:**
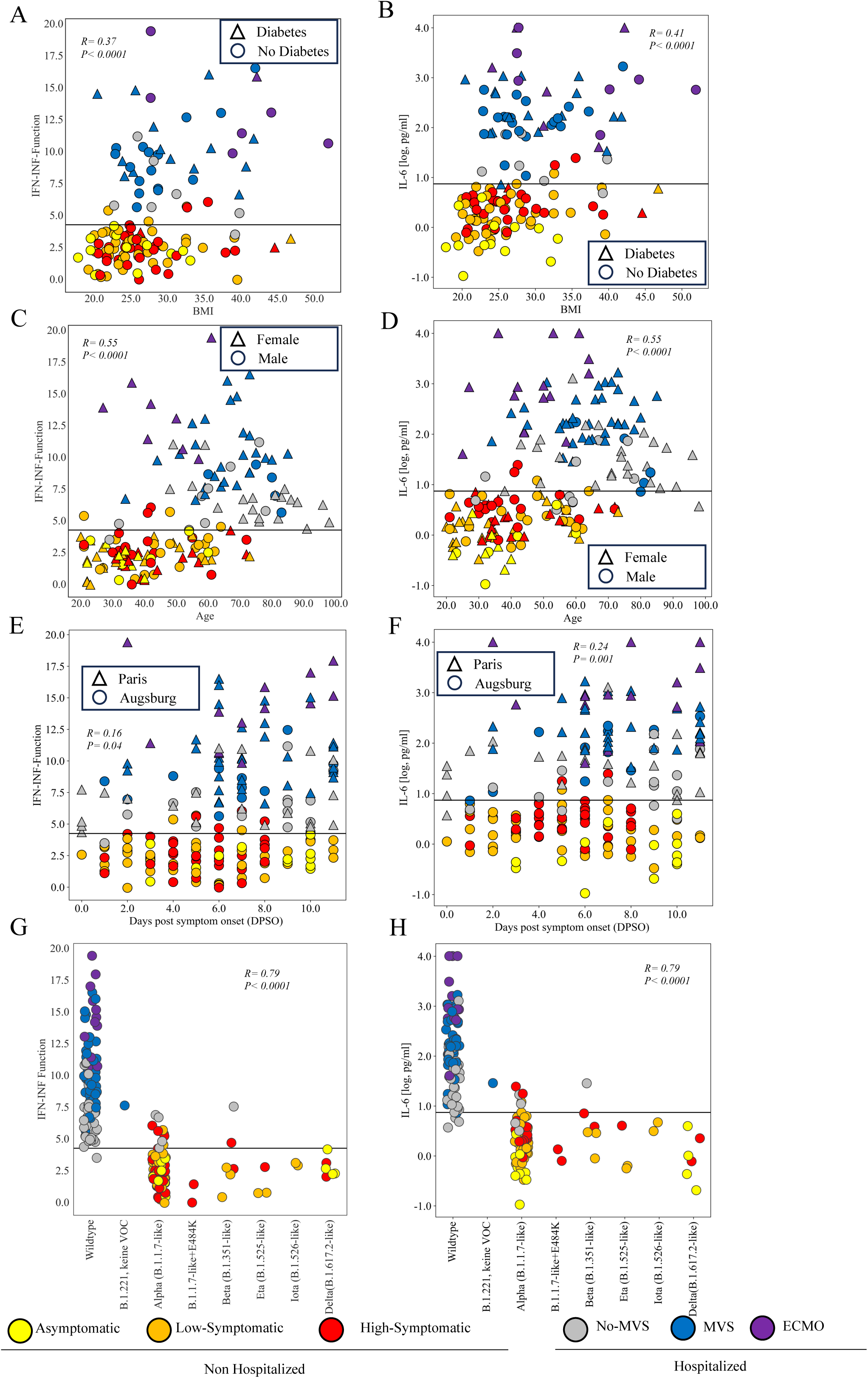
Independence of IFN-INF and IL-6 as predictors for hospitalization risk as function of different known risk factors for COVID-19 severity. Scatterplots show the independence from other factors of the prediction of hospitalization risk by IFN-INF (A, C, E and G) and IL-6 (B, D, F and H). Solid horizontal black lines represent the threshold for each biomarker for which the symptoms risk prediction has the largest Youden J-index (see Table 2). Although both IFN-INF and IL-6 are significntly correlated with BMI (A,B) and age (C,D), prediction by IFN-INF and IL-6 is independent of BMI (A,B), having diabetes (A,B), age (C,D), or gender (C,D). Importantly, no differences in the prediction power by IFN-INF or IL-6 are observed as function of time from symptom onset ranging 0-11 DPSO (E,F), or between the study sites (E, F). Lastly, the prediction of hospitalization risk by IFN-INF and IL-6 held for differrent SARS-CoV-2 variants (G,H). IFN-INF prediction function is defined by: log(2*IL-6*IL-10*TNF-α/IFN-α)

**Supplementary Figure 10:**
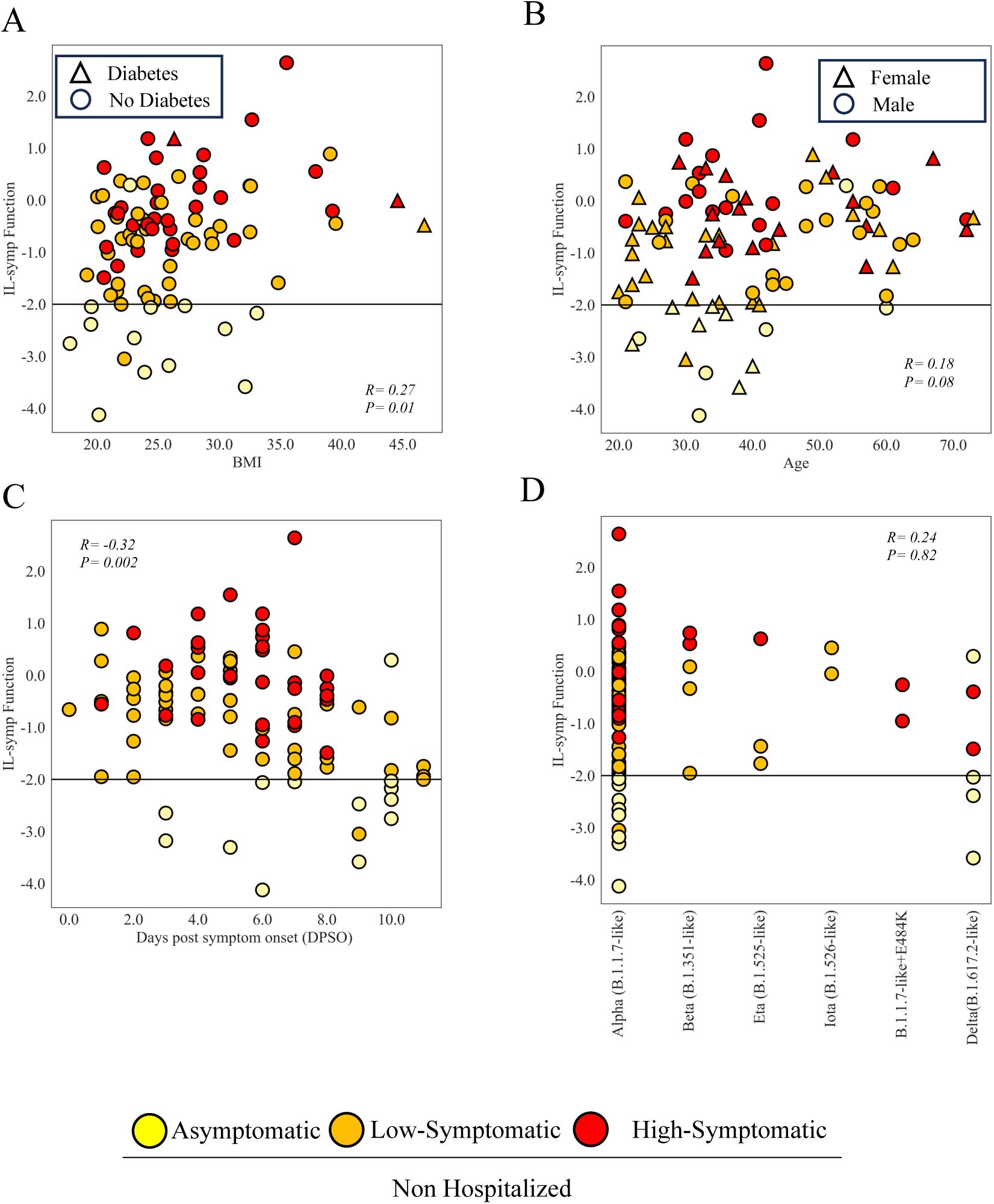
Independence of IL-symp as predictor for symptoms risk as function of different known risk factors for COVID-19 severity. Scatterplots show the independence from other factors of the prediction of symptoms risk by IL-symp. Solid horizontal black lines represent the IL-symp threshold for which the symptoms risk prediction has the largest Youden J-index (see Table 2). Prediction by IL-symp is independent of BMI (A), having diabetes (A), age (B), or gender (B). Importantly, no difference in the prediction power by IL-symp is observed as function of time from symptom onset ranging 0-11 DPSO (C). Lastly, the prediction of sympotms risk by IL-symp held for differrent SARS-CoV-2 variants (D). IL-symp function is defined by: -1.6*(log(IL-6)+0.175*log(IFN-γ))

## Notes

### Competing Interest Statement

The authors have declared no competing interest.

### Author Declarations

Ethics committee of the Technical University of Munich gave ethical approval for this work.

